# Psilocybin desynchronizes brain networks

**DOI:** 10.1101/2023.08.22.23294131

**Authors:** Joshua S Siegel, Subha Subramanian, Demetrius Perry, Benjamin Kay, Evan Gordon, Timothy Laumann, Rick Reneau, Caterina Gratton, Christine Horan, Nicholas Metcalf, Ravi Chacko, Julie Schweiger, Dean Wong, David Bender, Jonah Padawer-Curry, Charles Raison, Marcus Raichle, Eric J. Lenze, Abraham Z Snyder, Nico U.F. Dosenbach, Ginger Nicol

**Author notes:** equal contribution (Nico U.F. Dosenbach, Ginger Nicol).

## Abstract

The relationship between the acute effects of psychedelics and their persisting neurobiological and psychological effects is poorly understood. Here, we tracked brain changes with longitudinal precision functional mapping in healthy adults before, during, and for up to 3 weeks after oral psilocybin and methylphenidate (17 MRI visits per participant) and again 6+ months later. Psilocybin disrupted connectivity across cortical networks and subcortical structures, producing more than 3-fold greater acute changes in functional networks than methylphenidate. These changes were driven by desynchronization of brain activity across spatial scales (area, network, whole brain). Psilocybin-driven desynchronization was observed across association cortex but strongest in the default mode network (DMN), which is connected to the anterior hippocampus and thought to create our sense of self. Performing a perceptual task reduced psilocybin-induced network changes, suggesting a neurobiological basis for *grounding*, connecting with physical reality during psychedelic therapy. The acute brain effects of psilocybin are consistent with distortions of space-time and the self. Psilocybin induced persistent decrease in functional connectivity between the anterior hippocampus and cortex (and DMN in particular), lasting for weeks but normalizing after 6 months. Persistent suppression of hippocampal-DMN connectivity represents a candidate neuroanatomical and mechanistic correlate for psilocybin’s pro-plasticity and anti-depressant effects.

## 2 Main Text

Psychedelic drugs can reliably induce powerful changes in the perception of self, time and space via agonism of the serotonin 2A receptor (5-HT2A receptor)^1,2^. In clinical trials, a single high dose of psilocybin has demonstrates immediate and sustained symptom relief in depression^3–8^, and addiction^9^. Taken together, these two observations suggest that psychedelics should have distinct neurobiological signatures corresponding to acute (intoxication) and persisting (after the drug has been eliminated from the body) phases.

Acute effects of psilocybin include increase in glucose metabolism in frontal and medial temporal cortex^10,11^. EEG and resting state functional magnetic resonance imaging (rsfMRI) studies of acute psychedelic effects have reported decreased signal power^12,13^ and functional connectivity (FC) with psilocybin (Carhart-Harris et al., 2012), and LSD (Carhart-Harris et al., 2016) broadly across the cortex. These seemingly paradoxical observations of increased metabolism and decreased signal power are not understood.

Persisting effects of psychedelics include increase in the expression of genes that contribute to synaptic plasticity^14,15^, and increase in growth of neurites and synapses in vitro^16^, and in vivo^17–19^. Network-level correlates of persisting psilocybin effects remain unknown in humans.

Rodent models have suggested that the burst of plasticity in the medial frontal lobe and anterior hippocampus may be key to psilocybin’s antidepressant effects^16,17,20^. The medial frontal lobe and anterior hippocampus form core part of the default mode network^21^ and increased cortico-hippocampal connectivity has been associated with affective symptoms^22^.

Precision functional mapping (PFM) utilizes dense repeated rsfMRI sampling^23–25^ to reveal the timecourses of individual-specific intervention-driven brain connectivity changes^26^. The PFM approach takes advantage of the high variability in brain networks across individuals and high stability within individuals^24^. We applied the PFM approach to detail the individual-specific acute and persistent effects of a single high dose of psilocybin (25mg).

Healthy young adults received psilocybin (PSIL) and methylphenidate (MTP, generic Ritalin, dose-matched for arousal effects) on different days and underwent regular MRI sessions (∼18 per participant) each before, during, between, and after the two drug doses (Fig. S1A). Dense pre-drug sampling familiarized participants with the scanner and establish baseline variability. We found that psilocybin produced changes in brain network structure that far exceeded those induced by other state changes such as treatment with methylphenidate, or an auditory-visual task. Interrogation of the blood oxygenation level dependent (BOLD) signal revealed that the highly conserved spatial structure of resting BOLD fluctuations was dramatically altered by psilocybin. Finally, we identified a change in hippocampal-cortical connectivity that persisted for weeks after psilocybin and had normalized by the time participants returned for additional open label psilocybin doses 6+ months later.

### 2.1 Psilocybin massively disrupts networks

Psilocybin acutely caused profound and widespread brain network changes (Figs. 1a, S2, S3), greater than 3x larger than the effects of methylphenidate or other manipulations (task engagement = 1.30, methylphenidate = 1.51, high head motion = 1.46, psilocybin = 4.58, between person = 4.58). To put psilocybin’s effects in perspective, it helps to consider that the differences in an individuals’ brain organization on/off drug were as large as those between different persons (Fig. 1b).

**Figure 1.**
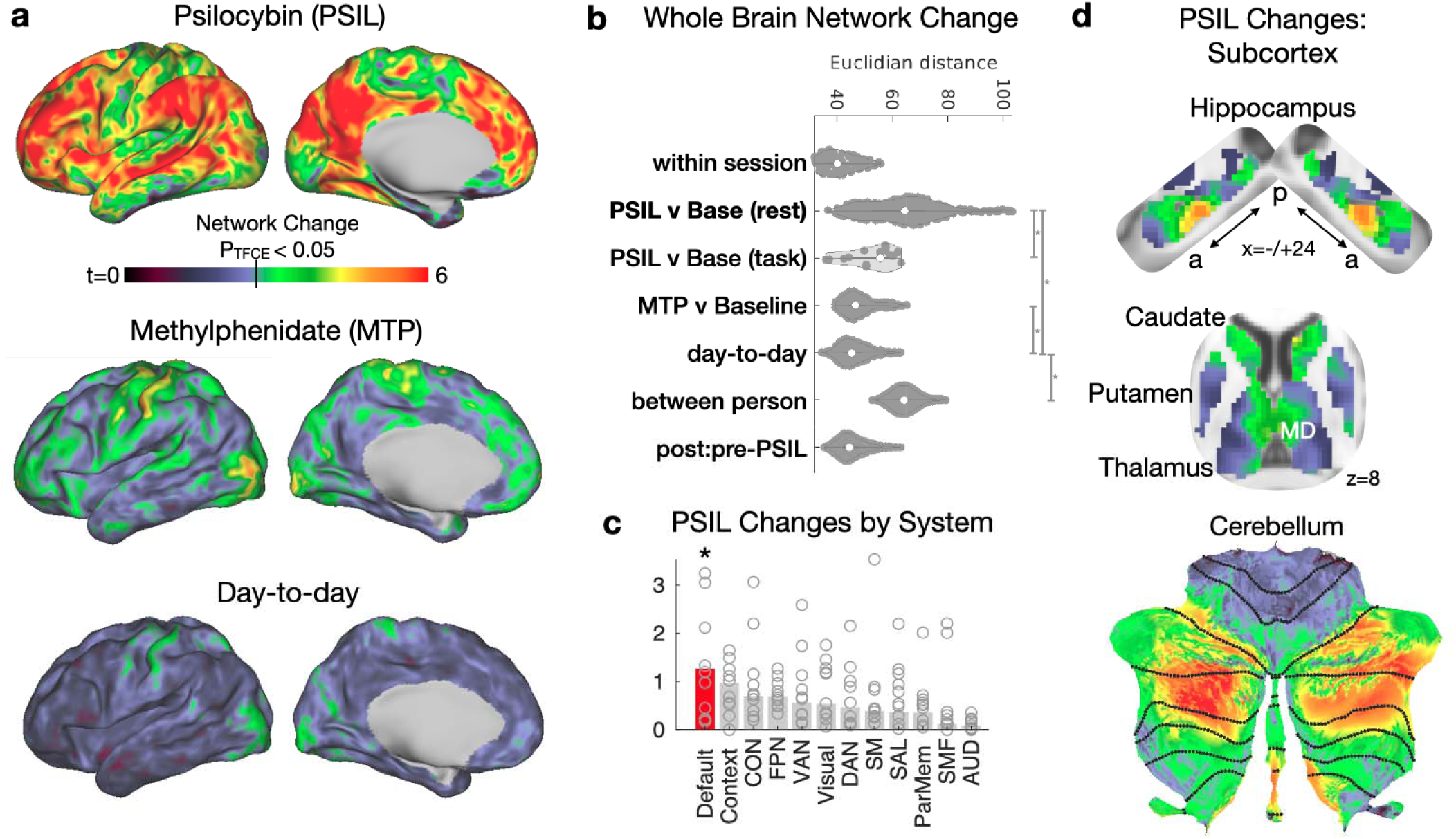
Psilocybin causes dramatic acute brain network changes. Brain-wide connectivity change (Euclidian distance) was calculated across the cortex and subcortical structures and effects of condition were determined with a linear mixed effects (LME) model. **a)** Top: psilocybin-associated network change (LME model, t-statistic). Middle: methylphenidate-associated network stimulant. Bottom: day to day FC variability (random effect of day). **b)** Whole Brain network change magnitude (Euclidian distance between brain-wide connectivity matrices) reveals that psilocybin produces much larger change than methylphenidate, closer in magnitude to between person differences. Vertical bars and asterisks indicate significance (p<0.05) for post-hoc t-test between conditions. **c)** Change by system (resting-state network), based on individualized Infomap parcellation. Open circles represent individual subjects, bars represent average magnitude. *Disruption was significantly enriched within the default mode network based on rotation permutation testing. d) Psilocybin Network Change across the bilateral hippocampus, basal ganglia, and cerebellum (same color scale as panel a).

In the cortex, psilocybin-associated changes localized to association networks. To assess resting state networks (RSNs, referred to here as ‘systems’) specificity, we ran a community detection algorithm on each participant and then compared network change between system, using a rotation-based null model^27^. Psilocybin-associated change was greatest in the default mode networks (Fig. 1c). By contrast, network changes associated with methylphenidate consistently localized to motor and action communities (Fig. S3). Individual-specific psilocybin-induced brain changes were consistent when participants returned 6 months later for a second psilocybin dose (correlation between change maps; r=0.67) (Fig. S2).

In subcortex, the largest psilocybin-associated network changes were seen in DMN-connected parts of the thalamus, basal ganglia, cerebellum, and hippocampus (Fig. 1d)^21,28,29^. In the hippocampus, foci of strong FC disruption were in the anterior hippocampus (MNI: -24,-22,-16 and 24,-18,16). In the basal ganglia, the largest FC disruptions were seen in mediodorsal (MD) thalamus and anteromedial caudate. In the cerebellum the largest FC disruptions were seen in the DMN^29^.

To examine the latent dimensions of variability in brain networks across participant, drug condition, and task, we performed multi-dimensional scaling (MDS) on brain networks from every session^24^. Dimension 1 (Dim1), the dimension which explained the largest amount of variability across the dataset, largely separated psilocybin from other scans (Fig. 2a), apart from one session where the participant (P5R) vomited 30 minutes after taking the psilocybin dose and was not re-dosed (red dots on the left of Fig. 2a). The higher score on Dim1 associated with psilocybin, corresponded to reduced segregation between default mode, frontoparietal, dorsal attention, and cingulo-opercular network – systems that are normally anticorrelated ^30^(Figs. 2c and S5). Subtraction of average FC during psilocybin minus baseline revealed a similar pattern of FC change (Fig. S5). To determine if this reflects a common effect of psilocybin that generalized across dataset and psychedelics, we calculated Dim1 scores for extant datasets from participants receiving I.V. psilocybin^31^ and LSD^32^. Psychedelic increased Dim1 in nearly every participant in the LSD and psilocybin datasets (Fig. 2d).

**Figure 2.**
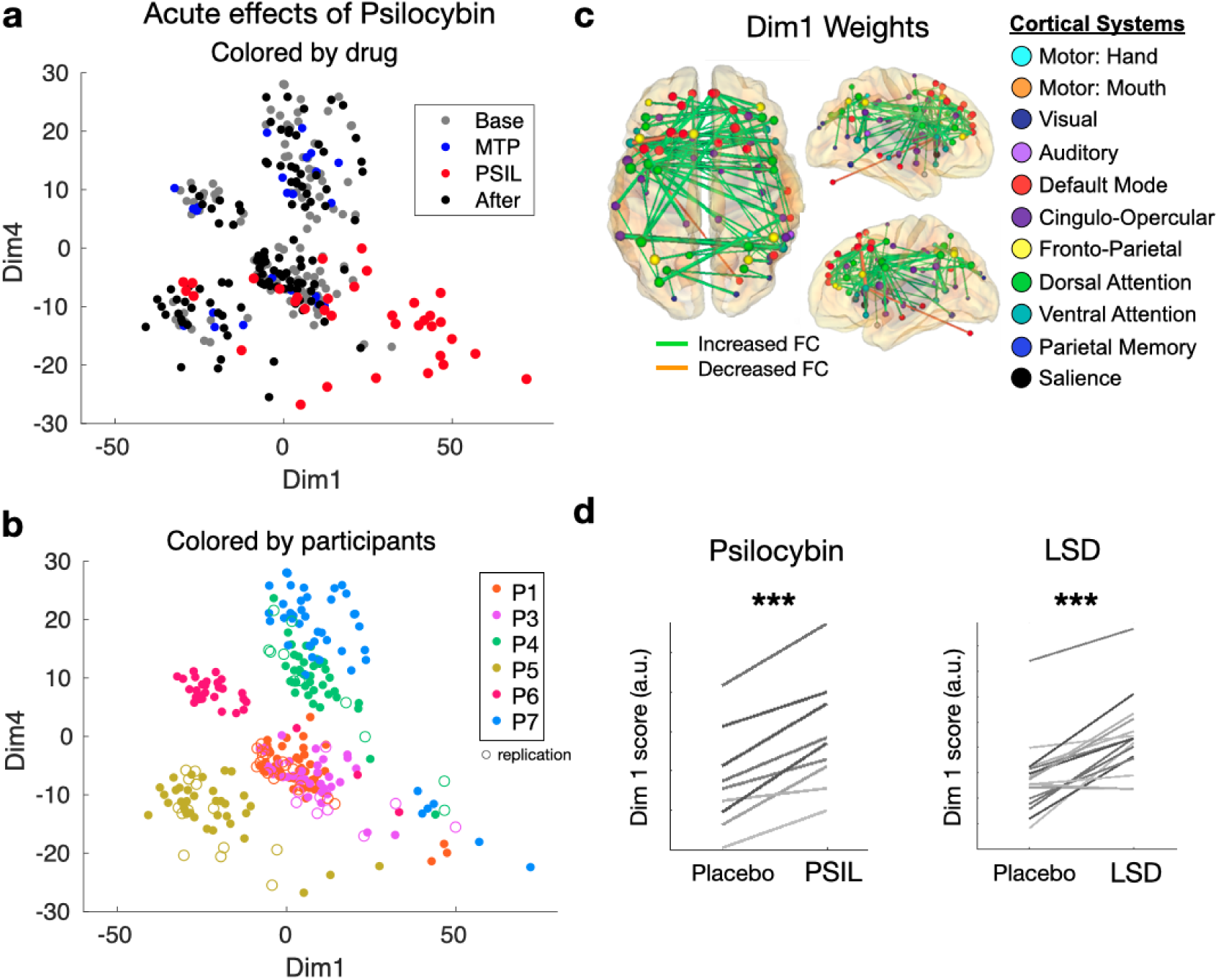
Multi-dimensional Scaling identifies acute and persistent drug effects. In the scatter plots, each point represents the functional network from a single scan, plotted in a multidimensional space based on the similarity between scans. Replication scans are represented with open circles. **a)** Dimensions 1 and 4 are plotted for every 15-minute scan. In the top panel, points are colored based on drug condition. In the bottom panel, points are colored based on participant identity. Dimension 1 separates psilocybin from non-drug and MTP scans in most cases. Dimension 4 separates individuals in most cases and shows a small but significant before/after effect. C) Visualization of dimension 1 weights (top 1% of edges are projected onto the brain to show the connections most effected by psilocybin). D) In additional datasets with LSD (top) and psilocybin (bottom), psychedelic causes reliable change along dimension 1.

By comparison, methylphenidate produced a decrease in connectivity between regions in the sensory, motor, and auditory networks (Fig. S5b), consistent with previous reports^33^ and similar to the effect of caffeine^25^. To verify that observations in our sample (N = 7) were generalizable, we compared our study’s stimulant effects to those in the Adolescent Brain Cognitive Development (ABCD) Study^34^ (n = 487 taking stimulants). The main effect of stimulant use on FC in ABCD was consistent with methylphenidate-associated FC changes in our dataset (Fig. S5b).

### 2.2 Disruption is mitigated by a perceptual task

To investigate how psilocybin-driven network changes are influenced by task states, participants were asked to complete a simple auditory-visual matching task in the scanner. Participants performed this task with high accuracy during drug sessions (Fig. S4). Engagement in the task significantly decreased the magnitude of psilocybin-associated network disruption by 43% (change _(psilocybin + rest)_ = 4.58; change _(psilocybin + task)_ = 2.59; unpaired t-test, P = 0.0041; Figs. 1b & S3c), suggesting that psilocybin’s acute effects on brain networks may be context dependent.

The reduction of psilocybin-driven brain changes parallels the psychological principle of ‘grounding’ – directing one’s attention on sensory perception as a means of alleviating intense or distressing thoughts or emotions – which is commonly employed in psychedelic-associated psychotherapy^35^. To our knowledge, this is the first evidence of context-dependent effects of psilocybin on functional connectivity and fills an important gap between preclinical studies of context dependence^36,37^ and clinical observations^38^.

Evoked responses during the auditory-visual matching task were largely unchanged during psilocybin, suggesting that altered neurovascular uncoupling is unlikely to account for observed FC change (Fig. S4).

### 2.3 Desynchronization at multiple spatial scales

We observed that the normally stable spatial structure of resting fMRI fluctuations was disrupted by psilocybin, making the FC desynchronized and disorganized (see video supplement). We measured synchrony of brain signals using Normalized Global Spatial Complexity (NGSC) – a measure of spatial entropy that is independent of the number of signals^39^. NGSC calculates cumulative variance explained by subsequent spatiotemporal patterns (Fig. 3a). The lowest value of NGSC=0 means that the timecourse for vertex/voxel is identical. The highest value of NGSC=1 means that the timecourse for every vertex/voxel is independent – indicating maximal desynchronization (or spatial entropy).

**Figure 3.**
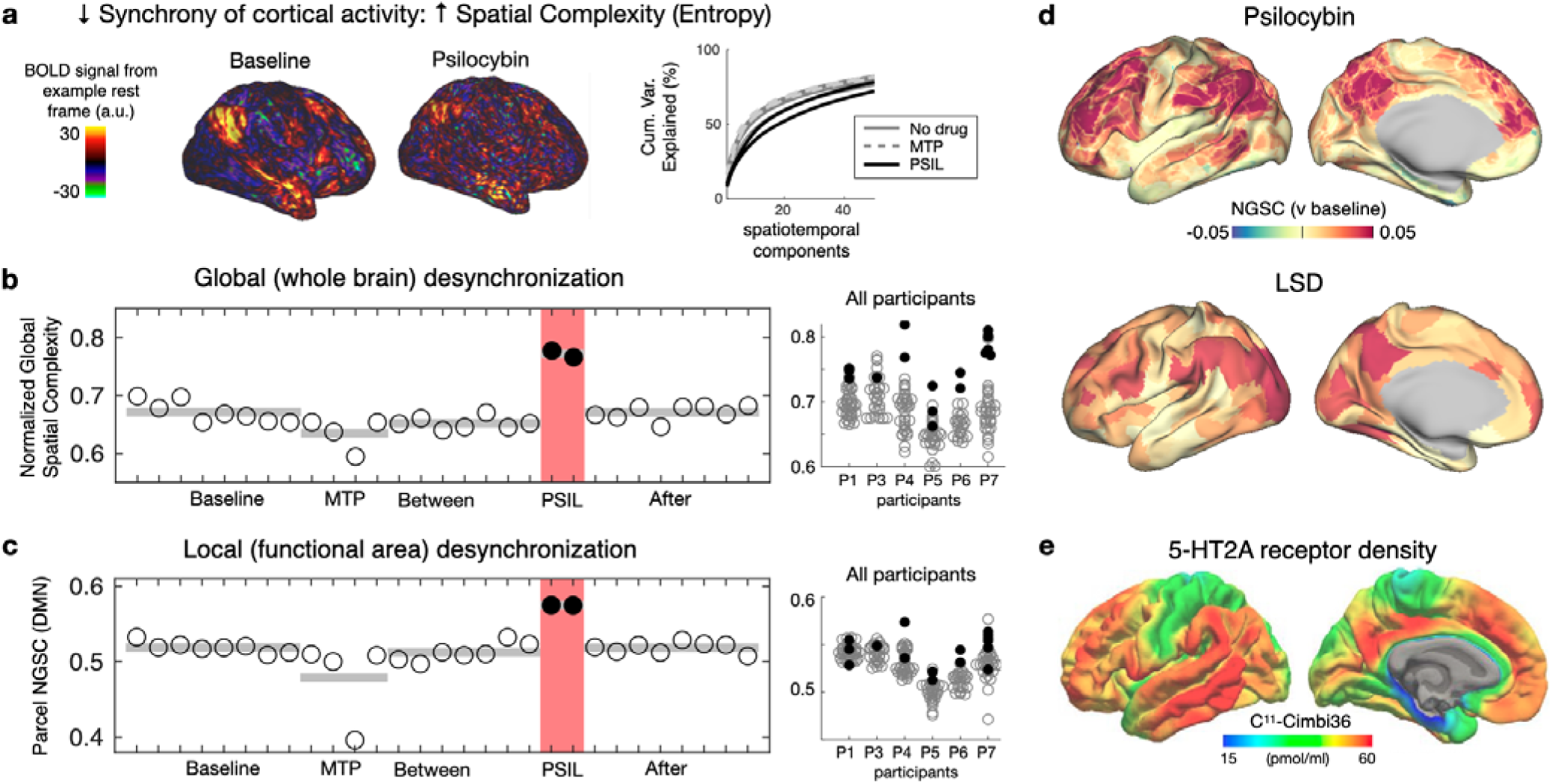
Increased spatial entropy (desynchronization of cortical activity) during psilocybin. **a)** normalized global spatial complexity (NGSC) captures the ‘complexity’ of brain activity patterns. It is derived from the number of spatial principal components (PCs) used to explain the data. Higher entropy = less desynchronized activity. **b)** Whole-brain entropy (NGSC) is shown for every fMRI scan for a single participant (P6). Psilocybin scans are shown in red. At right, increases during psilocybin were present in all participants (box and whiskers indicate quartiles and 99.7th percentiles for non-drug scans, red circles indicate psilocybin) **c)** Entropy within functional brain areas shown similar increases. **d)** Spatial entropy is visualized on the cortical surface. Psilocybin-associated increase in entropy was largest in association cortices. This was replicated in LSD data (middle; using data from Carhart-Harris et al., 2016) and corresponds spatially to 5ht2A binding density (bottom; borrowed with permission from Beliveau et al., 2017).

Psilocybin significantly increased NGSC (mixed effects model with individual differences and head motion as random effects, p = 9.2e-7), with levels returning to pre-drug baseline by the following session (Fig. 3b-d). The increase in NGSC was observed at whole-brain and individual brain area levels (Fig. 3c, d), with the largest increases in association cortex and minimal chance in primary cortex (Fig. 3d). Global and local desynchronization replicated in an LSD dataset (Fig. 3d)^32^ and the distribution of these effects closely matched PET-based maps of 5HT-2A receptor density (Fig. 3e)^40^.

### 2.4 Persistent decreases in hippocampal-DMN connectivity

To assess if persistent neurotrophic and psychological effects of psychedelics might be reflected in network changes weeks after psilocybin, we assessed network changes 1-21 days post-psilocybin (compared to pre-drug). Whole brain network change scores were small, and not significantly different from day-to-day variability, indicating that the brain’s structure had mostly returned to baseline (Fig. S2 and S3).

The anterior hippocampus forms part of the default mode network^21^ and atypical cortico-hippocampal connectivity has been associated with affective symptoms^22^. The same region of the anterior hippocampus which showed strong acute FC changes with psilocybin showed persistent connectivity alterations in the three weeks post-drug period (Fig. 4a and Supplemental figure 7). This persistent suppression of FC between the anterior hippocampus and cortex was found in all participants (Fig. 4b; LME model testing change in RMS AntHippo-cortical FC with head motion as a random effect: p = 5.1e-5).

**Figure 4.**
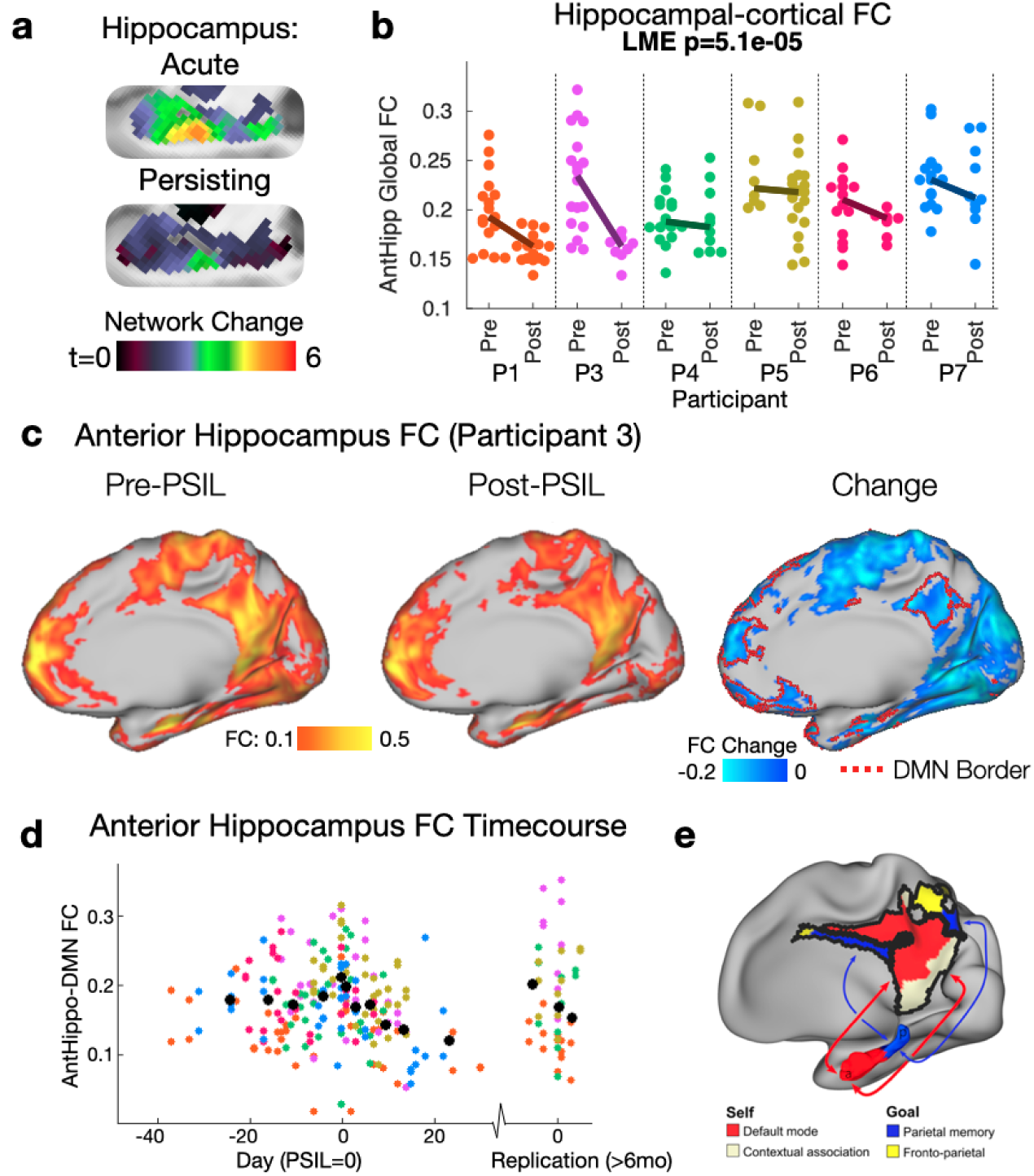
A persisting decrease in anterior hippocampal-DMN connectivity after psilocybin. **a)** Group network change (relative to baseline) t-maps, left hippocampus: top=acute psilocybin, bottom = persisting (2 weeks after psilocybin. **b)** All subjects demonstrated a weakening of anterior hippocampal-cortical FC post-psilocybin (p=5.1e-5; linear mixed effects model accounting for head motion and subject ID). **c)** Connectivity from an anterior hippocampus seedmap at baseline, post-psilocybin and change for an exemplar (P3). The red border on the right-most brain illustrates individual-defined default mode network. A decrease in hippocampal FC with parietal and frontal components of the DMN is seen. **d)** Timecourse of anterior hippocampus - DMN for all participants/scans (participant colors as in panel b). A moving average is shown in black. **e)** schematic of hippocampal-cortical circuits, borrowed from Zheng et al., 2021.

As predicted based on typical connectivity patterns, FC between the anterior hippocampus and DMN was decreased after psilocybin (Fig. 4c-e). Timecourse visualization, after aligning psilocybin doses between subjects, suggests that connectivity is reduced for 3 weeks after psilocybin but returned to pre-drug by 6–12-month replication scans (Fig. 4d). This observation is compelling, as it localized to the anterior hippocampus, a brain region shown to demonstrate substantial synaptogenesis following psilocybin^18^. Reduced hippocampal-cortical FC may reflect increased flexibility of self-oriented hippocampal circuits.

### 2.5 A link between acute and persisting effects

The synchronized patterns of co-activation in the resting brain are believed to reflect the brain’s perpetual job of modeling reality^41^. It follows that the stability of synchronized brain networks across day, task, methylphenidate, and arousal levels (but not between individuals), reflects the subjective stability of waking consciousness. By contrast, the much larger changes induced by psilocybin fit with participants’ subjective report of a radical change in consciousness. The large magnitude of psilocybin’s effects, in comparison to methylphenidate, suggests that observed changes are not merely a consequence of changes in arousal or a nonspecific effect of monoaminergic stimulation^42^.

Multi-unit recording studies suggest that agonism of 5HT2A receptors by psychedelics does not uniformly increase or decrease firing of pyramidal neurons, but rather serves to desynchronize pairs or populations of neurons which co-activate under normal conditions^43^. We observed a parallel phenomenon at the region, and whole brain level. This may explain the paradoxical effects of psychedelic seen in PET FDG and fMRI: disruption of synchronized activity should produce a decrease in power of local fluctuations^12,13^, despite an increase in metabolic activity, and a loss of the brain’s segregated network structure^32,44^. This desynchronization of neural activity has been described as an increase in entropy or randomness of brain activity in the psychedelic state^45,46^ (Carhart-Harris et al., 2014), and it has been hypothesized to underly the cognitive and perception changes associated with psychedelics.

The dramatic departure from normal synchronized patterns of co-activation may be key not only to understanding psilocybin’s acute effects, but also its persistent neurotrophic effects. Changes in resting BOLD activity are linked to shifts in glutamate-dependent signaling during psilocybin^47,48^. This phenomenon, shared by ketamine and psychedelics, engages homeostatic plasticity mechanisms^49^ - a neurobiological response to large deviations in typical network activity patterns^50–52^. This response to novelty includes rapid upregulation in expression of BDNF, mTOR, EF2 and other plasticity-related immediate early genes^15,48^. Consistent with this notion, psilocybin produced the largest changes in the fronto-parietal and default mode network, brain systems frequently associated with neuropsychiatric disorders^53^.

Psychedelics rapidly induce plasticity in the hippocampus and cortex and this appears to be necessary for rapid antidepressant-like effects in animal models. However, to understand the underpinnings of psychedelics’ unique effects, it is necessary to study the human brain. Advances in precision functional mapping and individual-level characterization enabled us to identify desynchronization of resting state fMRI signals and localize these changes to depression-relevant circuits. Novel methods to measure neurotrophic markers in the human brain^54^ will provide a critical link between mechanistic observations at the cellular, brain-systems and psychological levels. Such links will help answer fundamental questions about the diagnosis and treatment of conditions of psychiatric illness.

## 3 Methods

### 3.1 Regulatory approvals & registrations

Written informed consent was obtained from all participants in accordance with the Declaration of Helsinki and procedures established by the Washington University in Saint Louis Institutional Review Board. All participants were compensated for their time. All aspects of this study were approved by the Washington University School of Medicine (WUSM) Internal Review Board, the Washington University Human Research Protection Office (WU HRPO), the Federal Drug Administration (IND: 202002165) and the Missouri Drug Enforcement Agency (DEA) under a federal DEA schedule 1 research license and registered with clinicaltrials.gov (NCT04501653). Psilocybin was supplied by Usona Institute via Almac Clinical Services.

### 3.2 Study Design

Healthy young adults (N=7, 18-45 years) were enrolled in a randomized cross-over precision functional brain mapping study to evaluate differences in individual-level connectomics pre-, during, and post-psilocybin exposure. Participants underwent imaging during drug sessions with psilocybin (PSIL) 25mg, or methylphenidate (MTP) 40mg as well as non-drug imaging sessions. Drug condition categories were 1) before psilocybin (‘Baseline’), 2) Drug 1(methylphenidate or psilocybin), 3) between, 4) Drug2 and 5) After. A minimum of 3 non-drug imaging sessions were completed during each non-drug window: before (baseline), between and after drug sessions. The number of non-drug MRI sessions was dependent on availability of the participant, scanner and scanner support staff. Dosing day imaging sessions were conducted 60-180 minutes following drug administration during peak blood concentration^55^.

MTP was selected as the active control condition to simulate the cardiovascular effects and physiological arousal (i.e., controlling for dopaminergic effects) associated with psilocybin (Griffiths et al., 2006). Usona Institute, a United States non-profit medical research organization, provided good manufacturing practices (GMP) psilocybin.

Drug sessions were facilitated by two clinical research staff who completed an approved in-person or online facilitator training program provided by Usona Institute, as part of Usona’s phase 2 study (ClinicalTrials.gov identifier: NCT03866174). The role of the study facilitators was to build a therapeutic alliance with the participant throughout the study, prepare them for their drug dosing days, and to observe and maintain participant safety during dosing day visits^35^. The pair consisted of an experienced clinician (lead clinical facilitator) and a trainee (co-facilitator).

#### 3.2.1 Replication protocol

Participants were invited to return >6 months after completing the initial cross-over study for an open label psilocybin dose (which we refer to as ‘replication protocol). This included 1-2 baseline fMRIs, a psilocybin session (identical to initial session, except for blinding), and 1-2 “after” sessions within 4 days of the dose.

### 3.3 Participants

Healthy adults ages 18 to 45 years were recruited via campus-wide advertisement and colleague referral. Participants were enrolled from March 2021 to May 2023. Participants were required to have had at least one previous lifetime psychedelic exposure (e.g., psilocybin, mescaline, ayahuasca, LSD), but no psychedelics exposure within the past 6 months. Individuals with psychiatric illness (depression, psychosis, addiction) based on DSM-5 were excluded.

### 3.4 MRI

Participants were scanned roughly every other day over the course of the experiment (Fig. S1). Imaging was performed at a consistent time of day to minimize diurnal effects in functional connectivity ^56^. Neuroimaging was performed on a Siemens Prisma scanner (Siemens, Erlangen, Germany) in the neuroimaging labs (NIL) at the Washington University Medical Center.

Structural scans (T1w and T2w) were acquired for each participant at 0.9 mm isotropic resolution, with real-time motion correction. Structural scans from different sessions were averaged together for the purposes of Freesurfer segmentation and nonlinear atlas registrations.

To capture high resolution images of blood oxygenation level-dependent (BOLD) signal, we used an echo-planar imaging sequence with 2mm isotropic voxels, multi-band 6, multi-echo 5 (TEs: 14.20 ms, 38.93 ms, 63.66 ms, 88.39 ms, 113.12 ms), TR 1761ms, flip angle = 68 degrees, and in-plane acceleration^57^ (IPAT/grappa) = 2. This sequence acquired 72 axial slices (144mm coverage). Each resting scan included 510 frames (lasting 15:49 minutes) as well as 3 frames at the end used to provide estimate electronic noise.

Every session included two rest scans, totally 30-minute resting-state fMRI, during which participants were instructed to hold still and look at a white fixation crosshair presented on a black background. Head motion was tracked in real time using Framewise Integrated Real-time MRI Monitoring software (FIRMM)^58^. An eye-tracking camera (EyeLink, Ottawa) was used to monitor participants for drowsiness.

Participants also completed a previously validated event-related fMRI task initially designed for the purposes of modeling hemodynamic responses in the auditory, visual, and motor cortices. Two task fMRI runs were completed during a subset of imaging sessions. Task fMRI runs employed the same sequence using in resting fMRI, included 48 trials (24 congruent, 24 incongruent), and lasted a total of 410s. See supplemental methods ‘Matching task’ fMRI for details.

### 3.5 Resting fMRI processing and resting state network definition

Resting fMRI data were preprocessed using a in-house processing pipeline. Briefly, this included alignment, normalization, non-linear registration, removal of thermal noise using NORDIC^59^, bandpass filtering, and scrubbing at a movement threshold of 0.3mm. Tissue-based regressors were computed in volume (white matter, ventricles, extra-axial CSF)^60^ and applied following projection to surface. Details on rsfMRI preprocessing are provided in supplemental methods.

#### 3.5.1 Surface Generation and Brain Areal Parcellation

Surface generation and processing of functional data followed procedures similar to Glasser and colleagues 2013^61^. For cortical regions and resting state networks, we used a surface parcellation and community assignments generated by Gordon & Laumann and colleagues^27^. See supplemental methods for further details on surface generation and cortical parcellation.

For subcortical regions, we used a set of regions of interest (Seitzman et al., 2020) generated to achieves full coverage and optimal region homogeneity. A subcortical limbic system was defined based on neuroanatomy: amygdala, antero-medial thalamus, nucleus accumbens, anterior hippocampus, posterior hippocampus^62^. These regions were expanded to cover fill anatomical structures (e.g. anterior hippocampus)^63^.

To generate region-wise connectivity matrices, time courses of all surface vertices or subcortical voxels within a region were averaged. Functional connectivity (FC) was then computed between each region timeseries using Fisher z-transformed Pearson correlation.

#### 3.5.2 Individualized System Mapping

We identified canonical large-scale networks using the individual-specific network matching approach described in previously^27–29^. Briefly, cortical surface and subcortical volume assignemtns was derived using the graph-theory-based Infomap algorithm^64^. In this approach, we calculated the Pearson correlation matrix from all cortical vertices and subcortical voxels, concatenated across all a participant’s scans. Correlations between vertices within 30 mm of each other were set to zero. The Infomap algorithm was applied to each subject’s correlation matrix thresholded at a range of edge densities spanning from 0.01% to 2%. At each threshold, the algorithm returned community identities for each vertex and voxel. Communities were labeled by matching them at each threshold to a set of independent group average networks described in Gordon et al. 2016. In each individual and in the average, a “consensus” network assignment was derived by collapsing assignments across thresholds, giving each node the assignment it had at the sparsest possible threshold at which it was successfully assigned to one of the known group networks. See supplementary figure S2 and S3 for individual and group mode assignments.

### 3.6 “Network Change”, statistical approach

#### 3.6.1 Vertex-wise Network Change maps, approach and statistics

Network change (‘distance’) was calculated at the vertex level to generate network change maps (Fig. 1a-d, Fig. 4) and a linear mixed effects model with wild bootstrapping^65,66^ was used to generate statistics on these maps. First, a distance map was generated for every scan by computing, for each vertex, the average distance between its FC seedmap and the FC seedmap for each of that subject’s baseline scans. Second, distance maps for every scan were labeled on 5 dimensions: Subject ID, MRI session, task (task/rest), drug condition (baseline, PSIL, between, MTP, after, etc.), and head motion (average framewise displacement). Distance maps and associated conditions were then used to solve a linear mixed effects model in which drug condition and task were fixed effects, and Subject ID, MRI session, and head motion were random effects.

Wild bootstrapping in combination with threshold-free cluster enhancement (TFCE) was employed to estimate p-values for t-statistic maps resulting from the LME model. To perform the analysis, a wild bootstrapping procedure was implemented, which involved resampling clusters or groups of observations rather than individual data points. A large number of bootstrap samples (B=1,000) were generated using the Rademacher procedure^66^, where the residuals were randomly inverted to disrupt any potential autocorrelation. Specifically, a Rademacher vector was generated by randomly assigning -1 or 1 values with equal probability to each observation’s residual. By element-wise multiplication of the original residuals with the Rademacher vector, bootstrap samples were created to capture the variability in the data.

For each bootstrap sample, the TFCE algorithm was applied to enhance the sensitivity to clusters of significant voxels or regions while controlling for multiple comparisons. The value of the enhanced cluster statistic, derived from the bootstrap samples, was used to create a null distribution under the null hypothesis. By comparing the original observed cluster statistic with the null distribution, p-values were derived to quantify the statistical significance of the observed effect. The p-values were obtained based on the proportion of bootstrap samples that produced a maximum cluster statistic exceeding the observed cluster statistic. Significance thresholds were determined using alpha = 0.05 to control for the family-wise error rate.

The combined approach of wild bootstrapping with the Rademacher procedure and TFCE provided a robust and data-driven method to generate p-values for neuroimaging data. This methodology accounted for the complex correlation structure, effectively controlled for multiple comparisons, and accommodated potential autocorrelation in the residuals through the Rademacher procedure. By incorporating these techniques, significant effects of psilocybin and other conditions were reliably identified amidst noise and spatial dependencies.

#### 3.6.2 Whole Brain network distance

For analyses in Figs. 1b, 2, S2, S3, distance calculations were computed at the parcel level. The effects of day-to-day, drug condition (baseline, psilocybin, between, MTP, after), task, and head motion and their interactions were directly examined by calculating the distance between each functional network matrix. RMS Euclidean distance was computed between the linearized upper triangles of the parcellated FC matrix between each pair of 15-minute fMRI scans, creating a second-order ‘‘distance” matrix (Fig. S2). In this analysis, the two task sessions were concatenated to match the length of the 15-minute rest runs. Subsequently, the average distance was examined for functional network matrices that were (1) from the same individual within a single session, (2) from the same individual across days (‘‘day-to-day”), (3) from the same subject between drug and baseline (e.g. ‘‘PSIL v Base”), (4) from the same individual but different tasks (‘‘task:rest”), (5) from the same individual between highest motion scans and baseline (“hi:lo motion”), (6) from different individuals (“between person”).

Post-hoc comparison between drug conditions was conducted using two-tailed t tests. A related approach using z-transformed Pearson correlation (‘similarity’ rather than distance) was also taken and results were unchanged (Fig. S2c).

#### 3.6.3 Normalized Network Change

The different conditions above were compared by calculating whole brain network change magnitudes as shown in Figs. 1b, S2, and S3c. These are reported in the text as normalized network change scores calculated using the following procedure: We (i) determined network change for each condition compared to baseline as described above, (ii) subtracted within-session distance for all conditions (such that within-session network change = 0), (iii) divided all conditions by day-to-day distance (such that day-to-day network change = 1). Thus, normalized whole brain network change values (e.g. PSIL v Base = 4.58) could be thought of as proportional to day-to-day variability.

#### 3.6.4 Data-driven multidimensional scaling

We used a classical multidimensional scaling (MDS) approach to cluster brain networks across fMRI sessions, as described by Gratton and colleagues^24^. This data driven approach was used to identify how different parameters (e.g. task, drug, individual) affect similarity/distance between networks. MDS places data in multidimensional space based on the similarity (Euclidean distance; correlation-based distances produce similar results) among data points – where in this case a data point represents the linearized upper triangle of a given functional network matrix. Each separate matrix (from a given subject, task, and session) was entered into the classical MDS algorithm (implemented using MATLAB 2019, *cmdscale.m*). Multiple dimensions of the data were explored. The eigenvectors were multiplied by the original FC matrices to generate a matrix of eigen-weights that corresponded to each dimension.

#### 3.6.5 System specificity - rotation-based null model

To assess network specificity of vertex-wise PSIL network change values, we calculated average network change of matched null networks consisting of randomly rotated networks with preserved size, shape, and relative position to each other. Following Gordon & Laumann et al., 2016, we employed a rotation-based null model, in which a subject’s subnetworks were iteratively rotated a random amount around the spherical expansion of the cortical surface. The spatial autocorrelation of smoothed BOLD data means that small networks are more likely to contain a single connectivity pattern than large networks.

To create such matched random networks, we rotated each hemisphere of the original networks a random amount around the x, y, and z axes on the spherical expansion of the cortical surface. This procedure randomly relocated each network while maintaining networks’ size, shape, and relative positions to each other. Random rotation followed be computation of RSN-average PSIL network change score was repeated 1000 times to generate null distributions of network change scores. Vertices rotated into the medial wall were not included in the calculation. Actual PSIL network change score was then compared to null rotation permutations to generate a p-value for each of 12 systems (resting state networks) that were consistently present across every subject’s infomap parcellation.

For bar graph visualization of ‘PSIL Change by System’ (Figs. 1, S3), RSNs with greater change (p<0.05 based on null rotation permutations) are shown their respective color (as indicated in Fig. S3) and all other RSNs are shown in gray.

### 3.7 Normalized Global Spatial Complexity

We used a n approach previously validated to assess spatial complexity (termed entropy) or neural signals^39^. Temporal principal component analysis (PCA) was conducted on the full BOLD dense timeseries, which yielded m principal components (m = ∼80K surface vertices and subcortical voxels) and associated eigenvalues. The normalized eigenvalue of the i-th principal component was calculated as

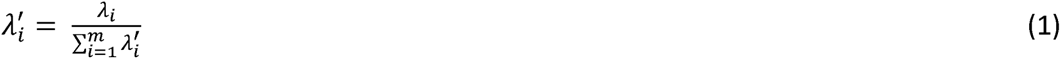

where m was the number of principal components, λ_i_ and λ′_i_ represented the eigenvalue and the normalized eigenvalue of the i-th principal component respectively. Lastly, the NGSC, defined as the normalized entropy of normalized eigenvalues, was computed using the equation:

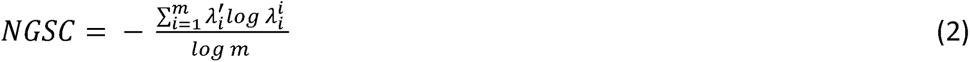

The NGSC computed above attains values from the interval 0 to 1. The lowest value NGSC = 0 would mean the brain-wide BOLD signal consisted of exactly one principal component or spatial mode, and there is maximum global functional connectivity between all vertices. The highest value NGSC = 1 would mean the total data variance is uniformly distributed across all m principal components, and a maximum spatial complexity or a lowest functional connectivity is found.

### 3.8 Persisting Effects Analysis

To assess if persistent effects of psychedelics, we compared network changes 1-21 days post-psilocybin to pre-drug baseline. The network change analysis (described above) indicated that connectivity at the ‘whole brain’ level did not change following psilocybin (Fig. S2 and S3). A screen was conducted with p>0.05 threshold to identify brain systems or areas showing persisting effects. This analysis identified the anterior hippocampus as a candidate region of interest (ROI) for persisting FC change (see ‘Pre/Post Psilocybin Network Change Analysis’ in Extended Data).

We assessed anterior hippocampus FC globally with cortex and specifically with the DMN. A bilateral anterior hippocampus ROI was used to generate a cortex-wide FC seed map for every session. Root-mean-square FC was computer to generate global FC^67^. Global FC was compared between pre-psilocybin sessions and post-psilocybin using a linear mixed effects model described above (drug condition and task were fixed effects, and Subject ID, MRI session, and head motion were random effects). Post-hoc visualization and timecourses were generated for FC with individual-defined default mode network.

### 3.9 Other Datasets

Raw fMRI and structural data from (Carhart-Harris et al., 2012) and (Carhart-Harris et al., 2016) were run through our in-house registration and processing pipeline described above. These datasets were used for replication, external validation, and generalization to another classic psychedelic (i.e., LSD) for the measures described above (e.g., normalized global spatial complexity, and the MDS-derived psilocybin FC dimension (Dimension 1).

Carhart-Harris et al., 2012: N=15 healthy adults completed two scanning sessions (psilocybin and saline) which included eyes-closed resting state BOLD scan for 6 minutes prior to and following I.V. infusion of drug. fMRI data were acquired using a gradient-echo EPI sequence, TR/TE 3000/35 ms, field-of-view = 192 mm, 64 × 64 acquisition matrix, parallel acceleration factor = 2, 90° flip angle.

Carhart-Harris et al., 2016: N=22 healthy adults completed two scanning sessions (LSD and saline), which included eyes-closed resting state BOLD scan acquired for 22 minutes following I.V. drug infusion lasting 12 min. fMRI data were acquired using a gradient echo planer imaging sequence, TR/TE = 2000/35ms, field-of-view = 220mm, 64 × 64 acquisition matrix, parallel acceleration factor = 2, 90° flip angle, 3.4mm isotropic voxels.

## Data Availability

De-identified data are available upon request.

---

**Table S1.** Participant demographics and neuropsychological assessments.

| Participant | P1 | P2 | P3 | P4 | P5 | P6 | P7 |
| --- | --- | --- | --- | --- | --- | --- | --- |
| **Basic demographics** |  |  |  |  |  |  |  |
| **Gender (n=3 females)** | Male | Male | Female | Female | Male | Female | Male |
| **Age Range (years)** | 41-45 | 36-40 | 36-40 | 36-40 | 18-20 | 21-25 | 41-45 |
| **Last degree completed** | Bachelor's | Bachelor's | Bachelor's | Graduate | High School | Bachelor's | Graduate |
| **Last psychedelic exposure (months)** | 24 | 24 | 12 | 24 | 24 | 12 | 60 |
| **Replication protocol (days between doses)** | Yes (273 days) | No | Yes (349 days) | Yes (350 days) | Yes (300 days) | No | No |
| **Mini-International Personality Item Pool (Mini-IPIP)** |  |  |  |  |  |  |  |
| **Neuroticism** | 3 | 2.5 | 2.75 | 2 | 2 | 1.25 | 2.5 |
| **Extraversion** | 2 | 3.5 | 3.25 | 3.25 | 4 | 3 | 3.5 |
| **Openness** | 2.5 | 3.75 | 1.75 | 4.75 | 4 | 5 | 3.75 |
| **Agreeableness** | 3.75 | 4.25 | 2.5 | 4.5 | 4 | 4.5 | 4.25 |
| **Conscientiousness** | 3 | 1.75 | 3 | 4.25 | 3.75 | 2.5 | 1.75 |
| **MRI data obtained** |  |  |  |  |  |  |  |
| **Number of usable 15-minute rsfMRI scans, without PSIL** | 53 | 44 | 40 | 44 | 41 | 26 | 17 |
| **Number of usable 15-minute rsfMRI scans, on PSIL** | 8 | 0 | 5 | 5 | 7 | 2 | 3 |
| **Total task MRI** | 9 | 9 | 7 | 9 | 3 | 6 | 16 |
| **Number of diffusion MRI, without PSIL** | 12 | 12 | 12 | 16 | 16 | 16 | 8 |
| **Number of diffusion MRI, on PSIL** | 2 | 0 | 2 | 2 | 2 | 0 | 0 |
| **Other protocol aspects** |  |  |  |  |  |  |  |
| **Respirations and pulse acquired** | Partial* | No | Partial* | Yes | Yes | Yes | Yes |
| **Completed replication protocol** | Yes | No | Yes | Yes | Yes | Yes | No |

**Figure S1.**
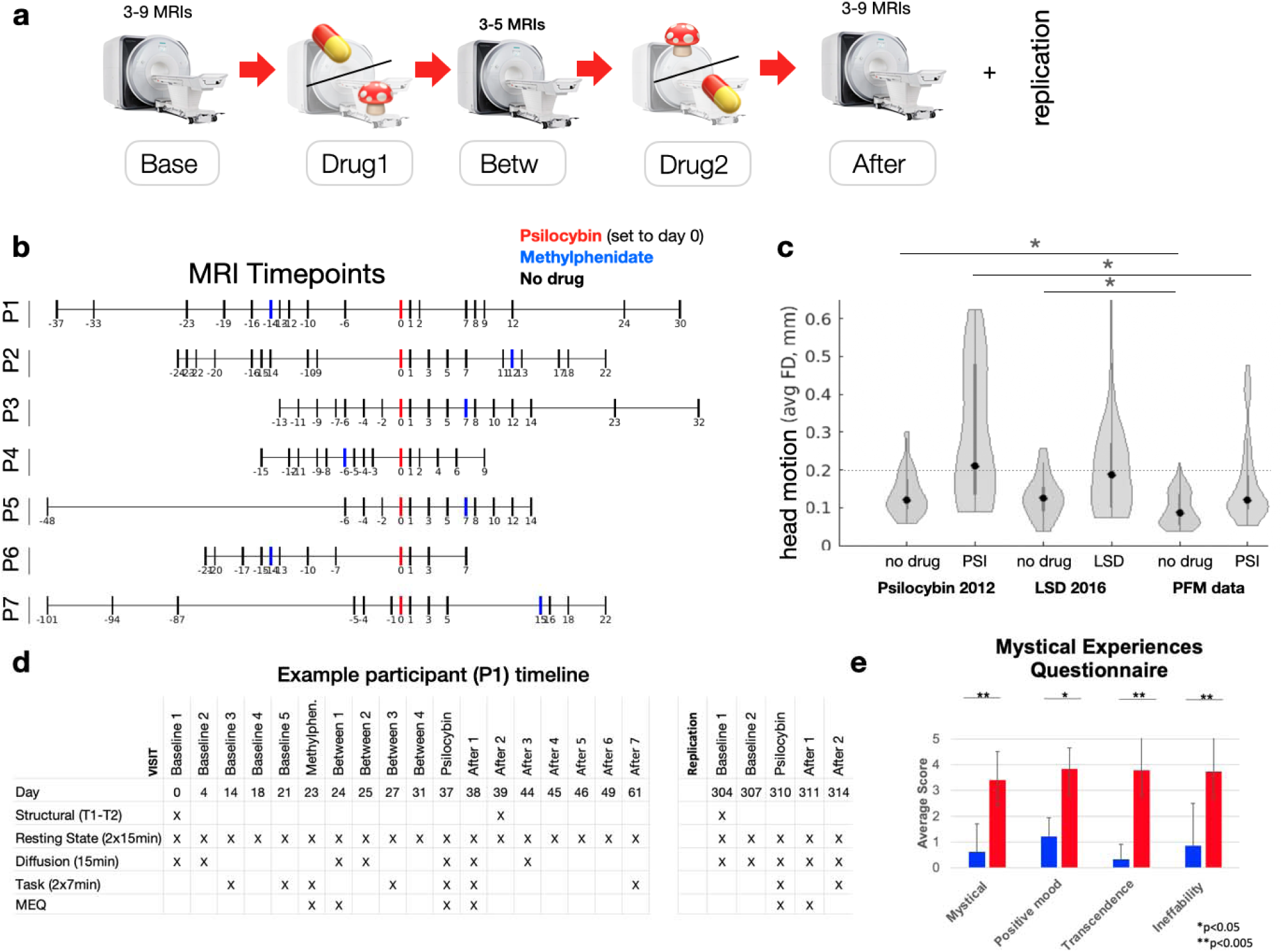
Quantifying Psilocybin effects with Precision Functional Mapping: design. **a)** Schematic illustrating the design of the precision functional mapping study of acute and persisting effects of psilocybin. Bringing participants in for multiple baseline visits enabled high-fidelity individual brain mapping, measurement of day-to-day variance, and acclimation to the scanner. **b)** Timeline of imaging visit for 7 subjects. **c)** Head motion comparison across datasets. Average head motion (FD, in mm) off and on drug is compared between our dataset and prior psychedelic fMRI studies (Carhart-Harris et al., 2012 & 2016). Dotted line at 0.2mm = recommended cutoff for usable fMRI scans. Asterisk: p<0.05, t-test. **d)** Timeline for an example participant. **e)** Participants reported significantly higher scores on all dimensions of the mystical experience questionnaire during psilocybin than placebo.

**Figure S2.**
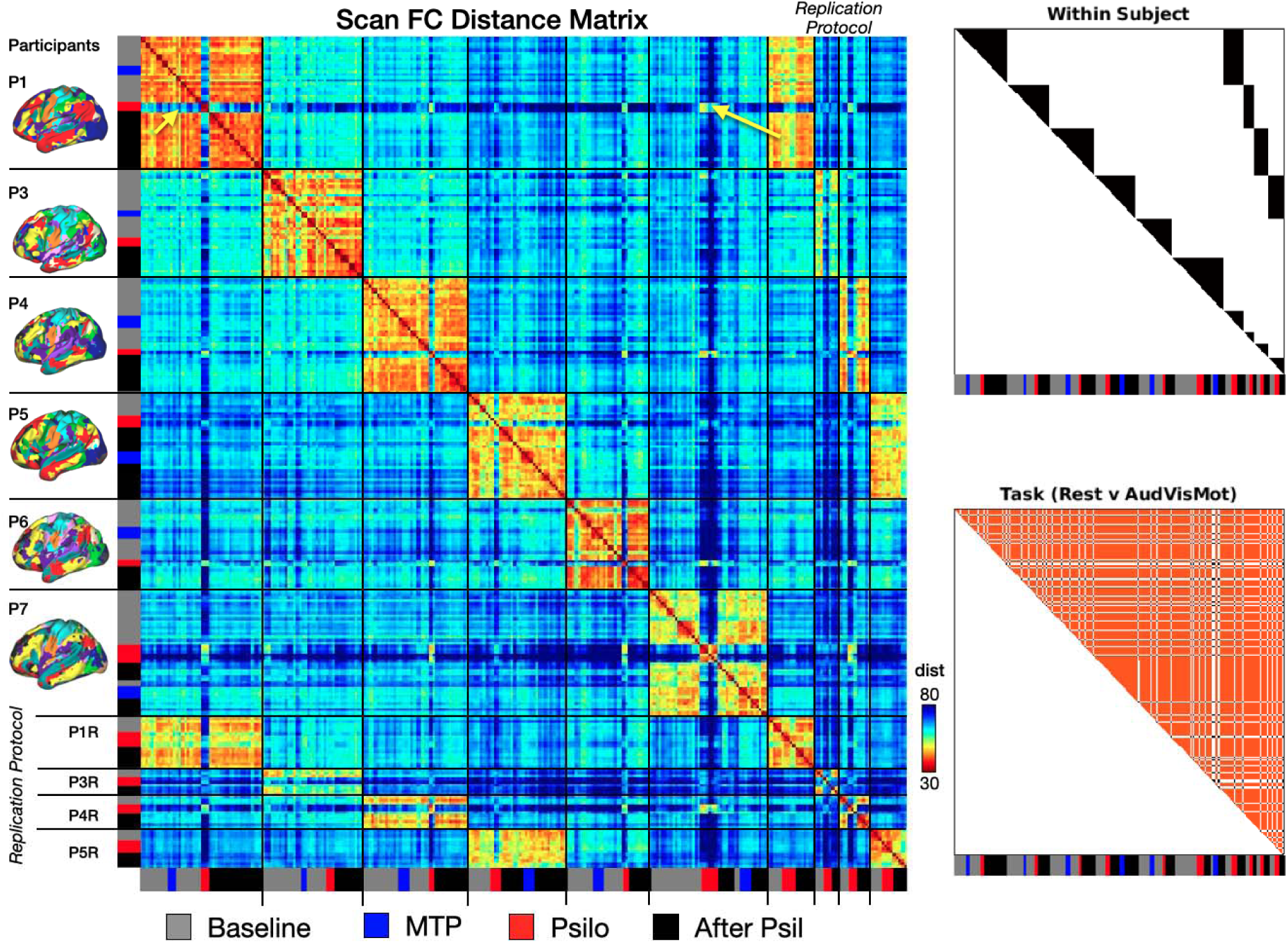
FC Distance and condition matrices. Following Gratton et al., 2018, we compare between rsfMRI sessions in order to quantify contributors to variability in functional brain networks. In this approach, the effects of group, individual, session, and drug (as well as their interactions) are examined by first calculating the Euclidian distance among every pair of functional network matrices (i.e., distance among the linearized upper triangles). LEFT: the resulting second-order ‘distance matrix’. Each row and column are brain networks from a single study visit. The values in the matrix indicate distance of functional networks between a pair of visits (i.e., Euclidean distance between the linearized upper triangles of two FC matrices). RIGHT: visualization of how the distance matrix was subdivided to compare different contributors to network change (typically relative to baseline scans). TOP: Black triages are separate subjects. Replication protocol visits are listed at the end. Note that psilocybin sessions (e.g. magenta arrow pointing to PS18 Psilocybin) are less similar to no-drug days, but more similar to psilocybin sessions from others, or in the same individual >6 months late

**Figure S3.**
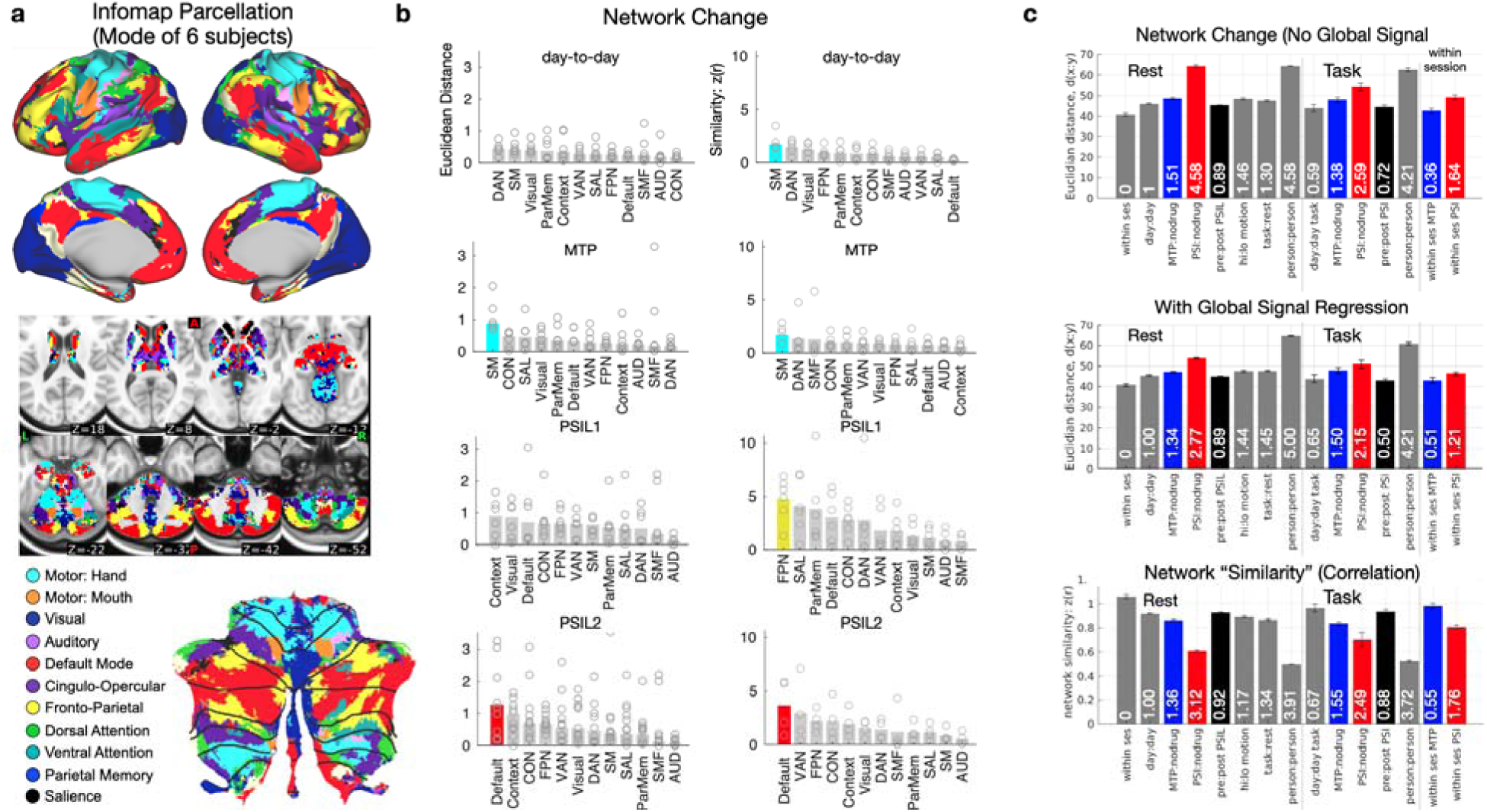
Network change compared across different conditions, brain structures, and measures. **a)** Mode Infomap-based RSN parcellations. **b)** Network selectivity of psilocybin-associated cortical change is assessed for different conditions (and separately for psilocybin initial and replication doses). Left column of bar plots shows network change based on Euclidean distance, right column is based on [decrease in] Pearson correlation. Colored bars indicates that the network showed change values that were above chance based on permutation of network labels (p<0.05, 10,000 null rotation). **c)** Network Change, defined as the average Euclidean distance between vectorized FC matrices, was examined before (top) and after (bottom) global signal regression that were (1) from the same individual within a single session, (2) from the same individual across days (‘‘day:day”), (3) from the same subject but different drug states (e.g. ‘‘psil:no-drug’’), (4) from the same individual but different tasks (‘‘task:rest”), (5) from the same individual between highest motion scans and baseline, (6) from different individuals (“person:person”). Bottom: Network change was also calculated using “similarity” (Pearson correlation) rather than difference, and yielded similar results.

**Figure S4.**
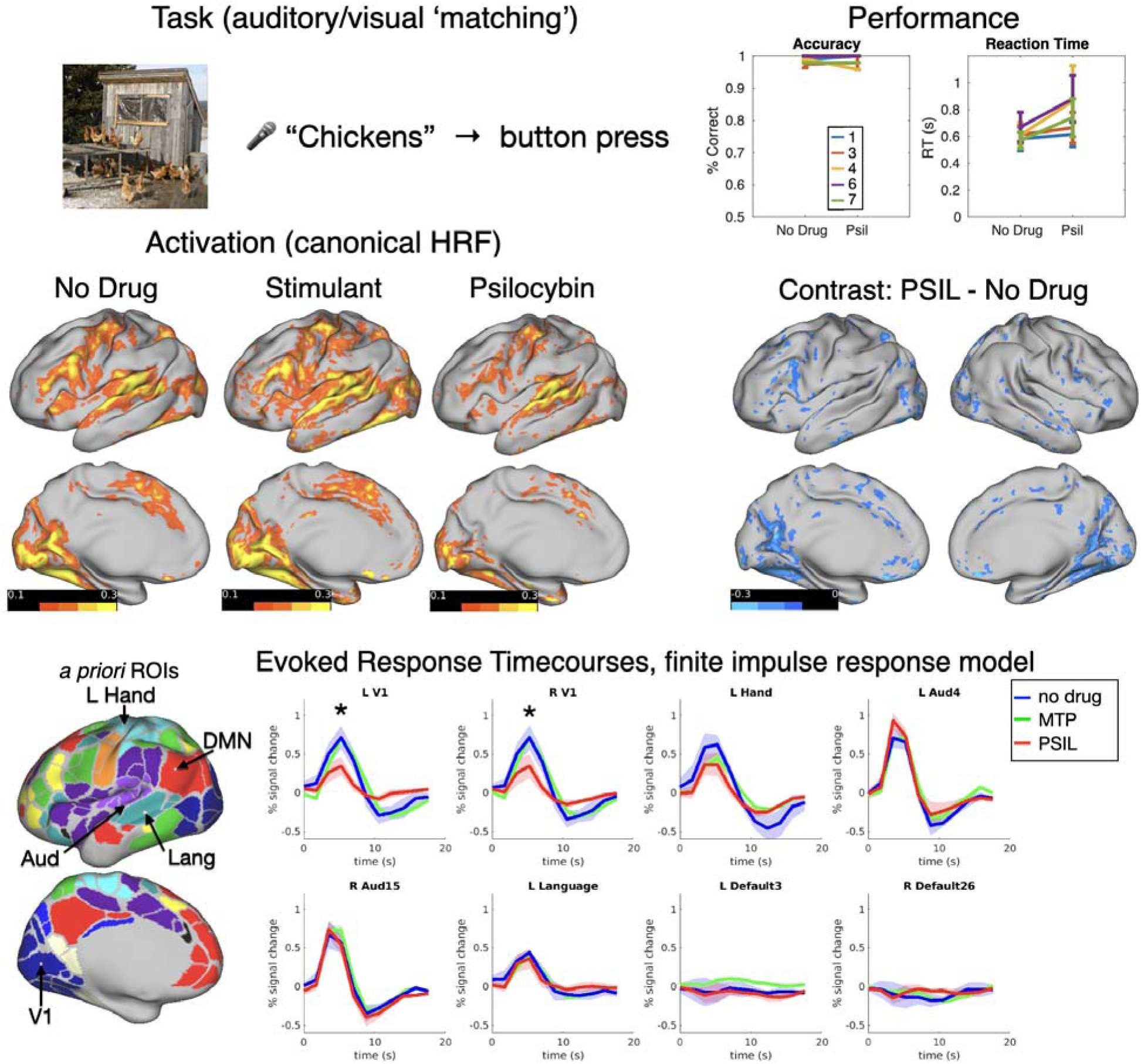
Auditory-Visual-Motor task. Top: Visualization of task design. Top right: psilocybin shows no effect on performance (both are at ceiling), but shows increase in RT latency and RT variability. Middle: Activation maps (left, beta weights) and contrasts (right, simple subtraction) using canonical HDR. Bottom: Average timecourses in 8 a priori regions of interest, calculated using FIR model. * P<0.05, ANOVA of Condition x HRF Betas (Main effect of all trials).

**Figure S5.**
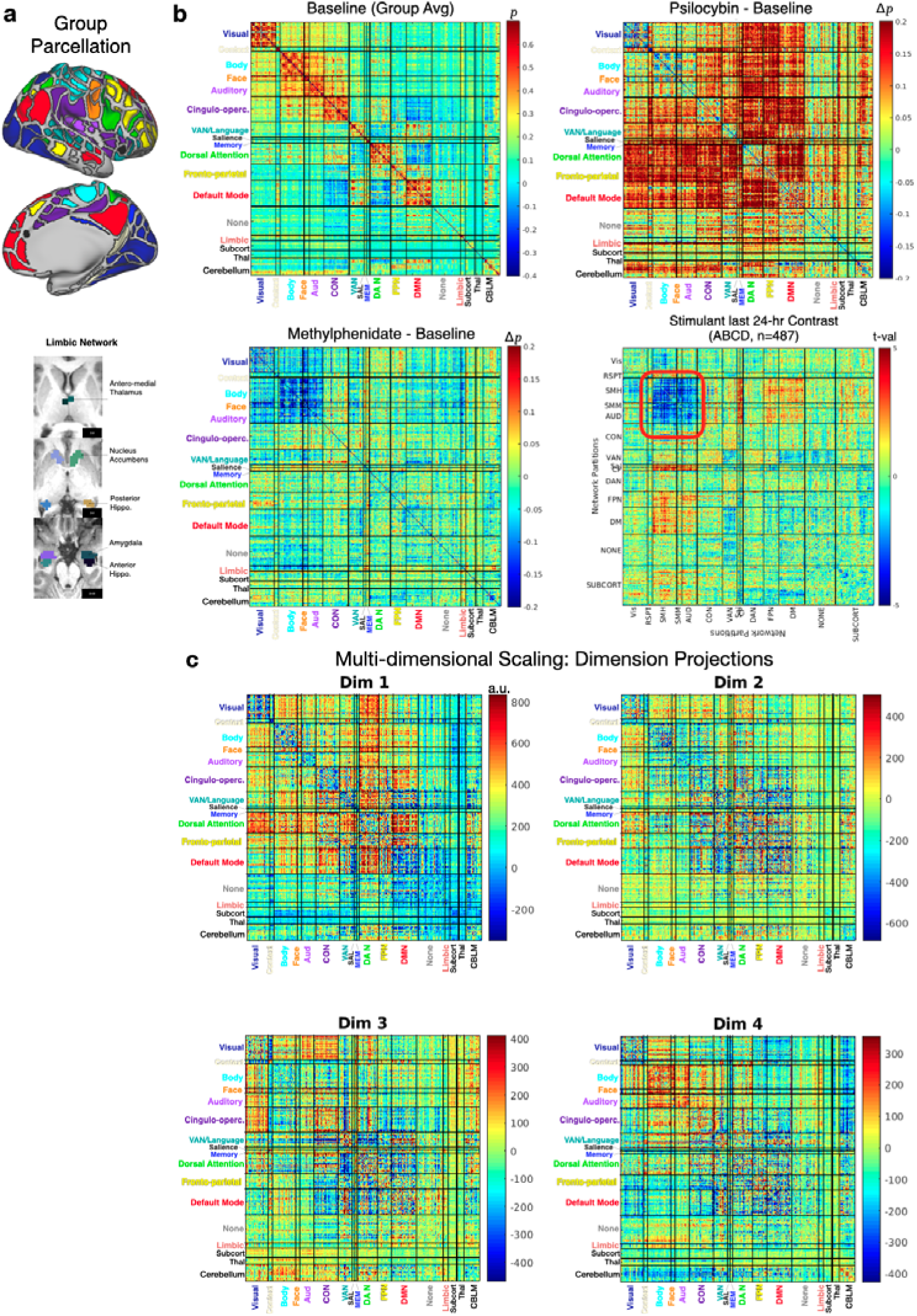
FC Matrices using Gordon-Laumann Parcellation. Top: Parcels and Average Condition FC Matrices. Top right: Psilocybin increases the correlation between Dorsal attention, Fronto-parietal, and Default Mode network to each other and to other cortical, limbic, and cerebellar systems. Top left: The group average FC ‘adjacency’ matrix, Bottom left: Methylphenidate minus baseline, Bottom right: for comparison and validation, we compared methylphenidate to the main effect of stimulant use within the last 24 hours (n=487 yes, n=8000 no) in ABCD fMRI data. B) Weights from the first 6 dimensions generated by multi-dimensional scaling of the full dataset. Dimension 1 shows strong acute psilocybin effect, dimension 4 shows weak pre-post psilocybin effect.

**Figure S6.**
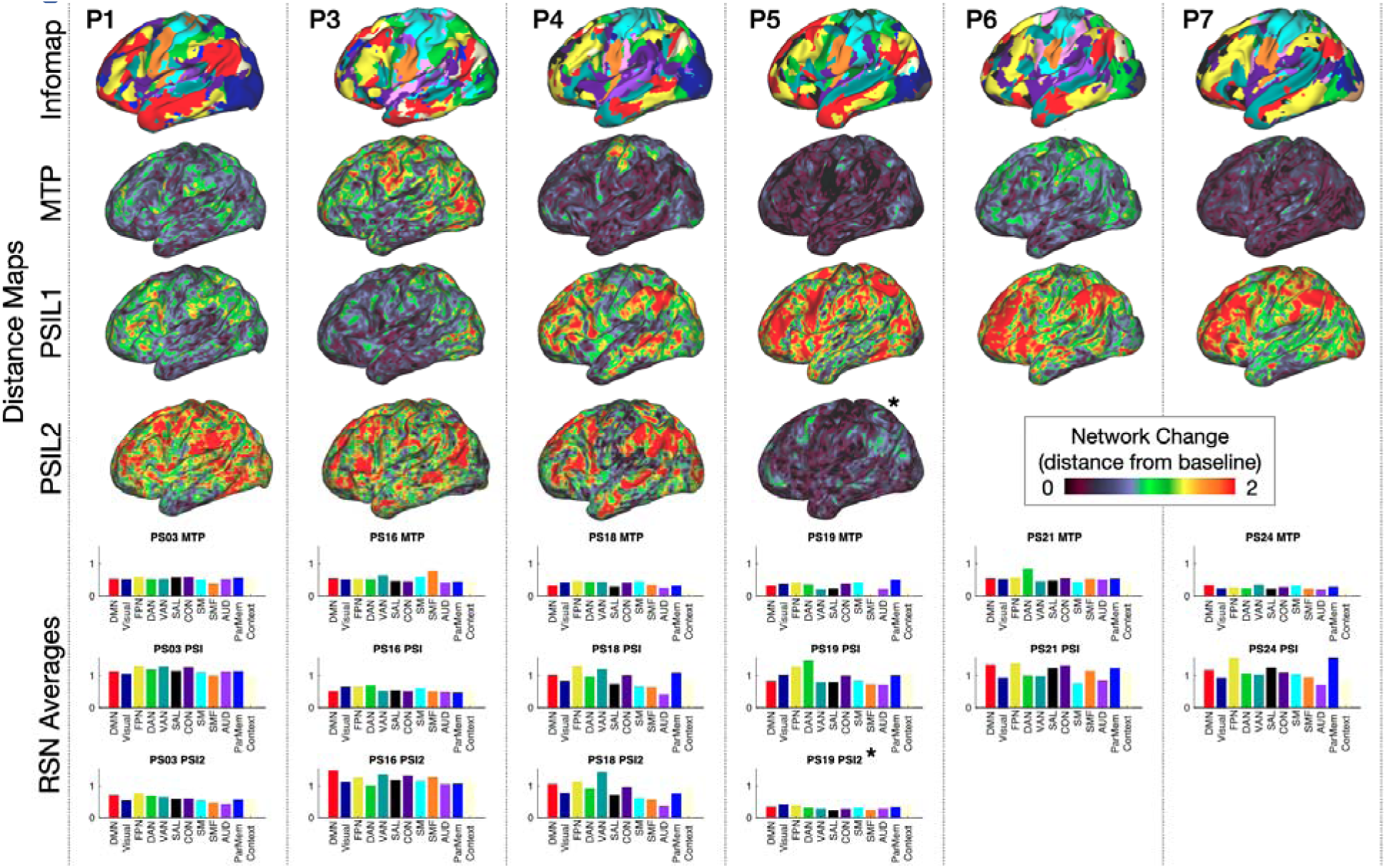
Individual subject MTP and PSIL Network Change maps. Top: Individual subject infomap parcellations. Middle: Network change maps, generated by calculating Euclidean distance from baseline seedmaps for each vertex. *Sub5 had an episode of emesis 30 minutes after drug ingestion during PSIL2. Bottom: Averaging distance maps within RSN generates RSN average network change scores (combined to map Fig. 1c).

**Figure S7.**
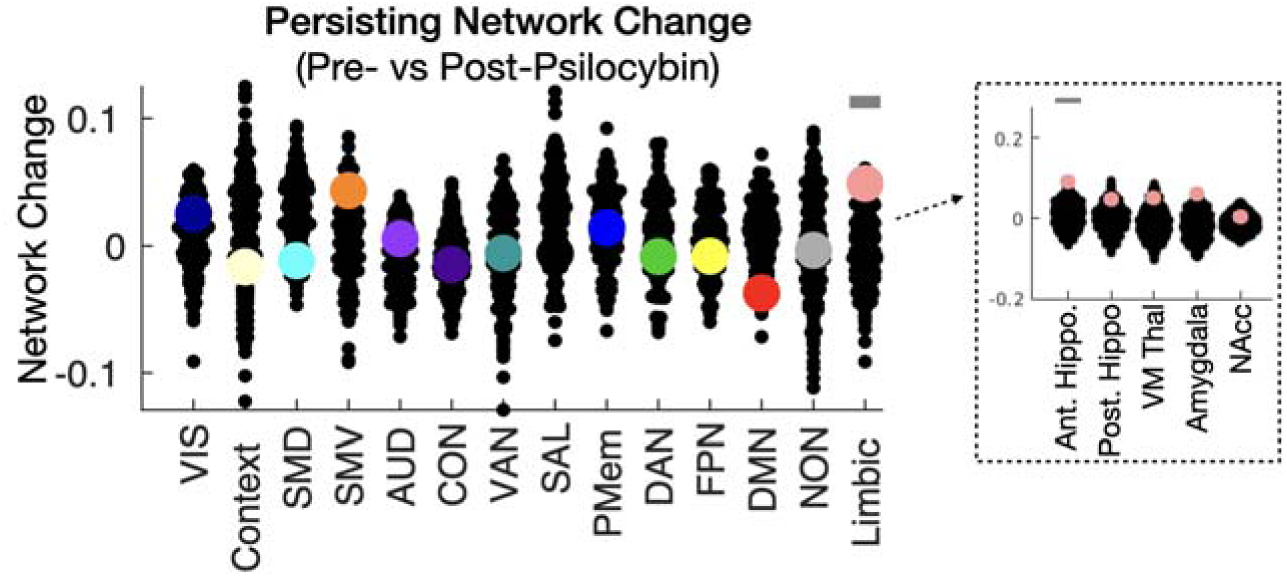
Pre/Post Psilocybin Network Change Analysis. Left: Permutation testing of persisting effects by system. Colored dots indicate network change for each system (baseline versus all post-psilocybin sessions). Black dots indicate network change for 500 permutations of pre/post labels. The gray bar above Limbic system indicates that persisting after psilocybin (p<0.05). Right: post-hoc analysis of the five bilateral regions of interest comprising the limbic system - anterior hippocampus, posterior hippocampus, ventromedial thalamus, amygdala, and nucleus accumbens. VIS = visual, SMD = somato-motor dorsal, SMV = somato-motor ventral, AUD = auditory, CON = cingulo-opercular network, VAN = ventral attention network, SAL = salience, PMem = parietal memory, DAN = dorsal attention network, FPN = fronto-parietal network, DMN = default mode network, NON = unassigned/low signal.

