## Supplemental Methods for "Psilocybin desynchronizes brain networks"

**Psilocybin profoundly alters brain networks**

***Supplemental Information***

### Exclusion criteria

Exclusion criteria included contradictions to MRI scanning (bone hardware, IUD, implantable devices) contraindications to psilocybin exposure (e.g., hypertension, cardiovascular disease, pregnancy); diagnosis of psychiatric condition (including substance use disorders); current use of certain psychotropic medication; previous adverse reactions to psychedelics; immediate family history of any schizophrenia spectrum disorder.

### Study screening

After prescreening and providing informed consent, participants underwent screening tests, including an electrocardiogram, urine drug screen, complete metabolic panel, and a urine pregnancy test. A study physician performed a physical exam and reviewed labs to ensure that participants did not have health conditions that would compromise their safety during the study. Once medically cleared, participants were scheduled for all planned imaging sessions and drug dosing days to ensure appropriate timing of pre-, post- and dosing day scans.

### Subjective and cognitive assessments

Assessments were conducted before, during, and after treatment sessions. This included subjective ratings, objective measures, personality survey and safety assessments. Assessments obtained are described below:

International Personality Item Pool-Five-Factor Model (Mini-IPIP): The Mini-IPIP is a 20-question survey administered to determine the Big Five factors of an individual’s personality: extraversion, agreeableness, conscientiousness, neuroticism, and openness to experience (Donnellan et al., 2006).  The Mini-IPIP was administered at the following time points: baseline, post-drug one, post-drug two.

Mystical Experience Questionnaire (MEQ): The MEQ is a 30-item self-report questionnaire that measures mystical experiences. It measures four factors: a) mystical (freedom from boundaries of one’s personal self and a feeling of unity to what is greater than one’s self), b) positive mood (sense of awesomeness or awe), c) transcendence of time and space (being outside of real of time), d) ineffability (sense that experience cannot be described well in words) (Barrett et al., 2015; Maclean et al., 2012). The MEQ was administered at the following time points: baseline, post-drug one, post-drug two. We administered the MEQ on the same day of drug administration and the day after dosing day and determined that MEQ scores were the similar on both days. Therefore, MEQ scores on dosing days were reported.

### Set and setting protocol

Preparation and integration sessions were held in a dedicated research treatment room where the study drug was administered. Preparatory sessions were held one or two days before drug administration. Integration sessions were held one day after drug administration. The purpose of preparatory sessions was to build a therapeutic alliance between facilitators and participants. The participant’s personal history, developmental stage, current life situation, and intentions for and expectations of drug sessions were reviewed. Preparation and integration sessions occurred per Usona facilitator training guidelines.

### Drug administration

On dosing visits, following checking vitals, urine drug screen, and urine pregnancy test, participants received either 25mg of psilocybin or 40mg of methylphenidate. Both facilitators and subjects were blinded. Medications were taken with lemon ginger tea. Following a 10 minute guided mindfulness meditation, participants were invited to lie on the sofa with eye shades as well as headphones and a curated music play list. One hour after drug administration, participants were transported to the MRI suite. Following the MRI, participants were transported back to the dedicated testing room and encouraged to direct their attention internally until subjective drug effects were resolved. When drug effects were resolved, study facilitators and participants completed post-dose questionnaires and a release checklist. Regardless of drug received, dosing sessions were 6-8 hours in length.

Heart rate and blood pressure were measured at regular intervals during dosing days (e.g., 30, 60, 90, 120, 240, 360, 420, and 480 minutes after drug ingestion). Subjects were also briefly queried about adverse effects during vital signs monitoring using an adverse events checklist. Rescue medications (risperidone for agitation, lorazepam for anxiety and niacin for chest pain) were available as needed. The Columbia Suicide Severity Rating Scale (C-SSRS) was used to assess for suicidal ideation and behavior during drug exposure (Posner et al., 2007). Participants had access to a physician who was physically present throughout dosing day.

### Treatment guess

After each drug session, participants were asked to guess if they had received psilocybin or MTP.

**Data management**

De-identified assessment scores, raw data from structural MRI and fMRI scans was uploaded into the Central Neuroimaging Data Archive (CNDA).

### Resting-State Functional MRI Processing and Surface Projection

Preprocessing of fMRI data included: 1) compensation for asynchronous slice acquisition using sinc interpolation; 2) elimination of odd/even slice intensity differences resulting from interleaved acquisition; 3) whole brain intensity normalization to achieve a mode value of 1000; 4) removal of distortion using field map and spatial realignment within and across fMRI runs; 5) resampling to 2mm cubic voxels in atlas space including nonlinear realignment and atlas transformation in one resampling step. Cross-modal (e.g., T2-weighted to T1-weighted) image registration was accomplished by aligning image gradients (2); 6) removal of thermal noise using NORDIC (a local PCA approach in which temporal components of an fMRI signal that are indistinguishable from Gaussian noise are eliminated) (3).

Following cross-modal registration, data were passed through several additional preprocessing steps: (i) tissue-based regressors were computed based on FreeSurfer segmentation (4); (ii) removal by regression of the following signals that contain spurious variance: (a) six parameters obtained by rigid body correction of head motion, (b) signal from white matter, ventricles and extra-axial sources of noise; (ii) temporal filtering to retain frequencies in the 0.009–0.08-Hz band; and (iii) frame censoring. Where indicated, respiratory and pulse-ox traces were used to generate additional physiological regressors using the PhysIO software package (5).

The first four frames of each BOLD run were excluded. As has been reported previously, several subjects exhibited high-frequency peaks in the power spectrum of head motion time courses, primarily in the phase-encoding (y) dimension. Thus, we low-pass filtered the motion time courses at 0.1Hz in all subjects prior to computing FD to prevent superfluous data loss (6,7). Frame censoring was implemented using framewise displacement (8) with a threshold of 0.3 mm. This frame-censoring criterion was uniformly applied to all rsfMRI data before functional connectivity computations. BOLD runs were excluded completely if they retained less than 50% usable frames after motion scrubbing.

Individualized cortical surfaces and subcortical volumes were generated for each subject’s T1 MRI using FreeSurfer automated segmentation (4). Segmentation errors were manually corrected. Following preprocessing, BOLD data were sampled to each subject’s individual cortical surface and subcortical volume using Connectome Workbench (9).

Brain surface visualizations were generated using Connectome Workbench (9).

### Individualized parcellation and Infomap Network Detection

### Brain Areal Parcellation

For cortical regions and resting state networks, we used a surface parcellation and community assignments generated by Gordon & Laumann and colleagues (10). Based on that parcellation (Figure S1), the following cortical resting-state FC networks were defined using infomap: Salience (black); Visual network (blue); Default Mode Network (red); Dorsal Attention (green); Ventral Attention/Language (teal); Congulo-opercular (purple); dorsal somato-motor (sky blue); ventral somato-motor (orange); auditory (pink); frontoparietal network (yellow); Retrosplenial-temporal (RSPT; white); Cingulo-parietal (cerulean); Low Signal (gray, outline in black). The Low Signal regions were excluded from analysis because they occur in known areas of signal dropout. Given their location (such as orbital frontal cortex and temporal pole) these regions

### Pre/Post Psilocybin Network Change Analysis (system level)

Pre/Post Psilocybin Network Change was computed at the system level for 13 cortical systems plus the limbic system. Permutation testing was employed to assess the statistical significance of the network change scores between pre- and post-drug conditions for each system. This procedure involved shuffling the labels of the pre- and post-drug session labels (while maintaining session-level and subject-level structure). Specifically, 500 permutations of the pre- and post-drug labels were generated, randomly reassigning the labels across sessions. For each permutation, the difference in network change scores between the shuffled pre- and post-drug conditions was calculated. This generated a distribution of differences expected under the null hypothesis of no difference between pre- and post-drug conditions.

Next, the observed difference in network change scores between the actual pre- and post-drug conditions was compared to the null distribution. The proportion of permuted samples that produced a difference greater than or equal to the observed difference provided the empirical p-value for assessing the significance of the observed difference. If the observed difference fell within the extreme tails of the null distribution, it was considered statistically significant, indicating a significant difference between pre- and post-drug network change scores for a given system.

This analysis indicated that the limbic system was the only system to show network change after psilocybin that exceeded chance (p-value < 0.05, uncorrected). Running the same analysis pre/post methylphenidate did not identify any systems with network change surpassing p < 0.05. Following this observation, we probed network change in the constituent parts of the limbic system. We separated the limbic system into five bilateral regions of interest - anterior hippocampus, posterior hippocampus, ventromedial thalamus, amygdala, and nucleus accumbens – and repeated the network change permutation testing approach 500 times. Here, the anterior hippocampus was the only region for which pre- and post-drug network change exceeded all 500 label permutations.

### Matching task fMRI Design

This was a suprathreshold auditory-visual matching task in which participants are presented with a naturalistic visual image (duration 500 ms) and coincident spoken English phrase and are asked to respond with a button press to indicate of the image and phrase are ‘congruent’ (for example, an image of a beach, and the spoken word beach) or ‘incongruent’. Both accuracy and response time of button press were recorded. Each trial was followed by a jittered inter-stimulus interval optimized for event-related designs. Each task fMRI session included two blocks, each lasting 410sec and including 48 trials (24 congruent, 24 incongruent),). On average, participants completed the task fMRI paradigm on 7.2 visits each.

### Task fMRI Analysis

fMRI data were preprocessed similar to resting data, with the exception of no nuisance regression and an FD threshold of 0.7mm (11). A generalized linear model was created by convolving 2x2 design matrix (congruent x incongruent, button press x no press) with an ~18s finite impulse response model. This allowed for possible changes to the shape of the hemodynamic response function on psilocybin. Additional demean and detrend terms were added to the generate a general linear model (GLM). This GLM was solved to estimate beta weights separately for each task run. In level 2 analysis, beta weights were averaged for all runs in a session and z-statistics were calculated.

A set of a priori regions of interest (ROIs) relevant to the task were selected from the Gordon-laumann parcellation. These included: left/right calcarine sulcus (V1), left/right auditory cortex (A1), left language (Wernicke’s area), left hand knob, left angular gyrus, and right angular gyrus (default mode).

Main effect of stimulus (z statistics) was compared between conditions using a multi-way ANOVA (matlab: anovan) with subjects as a random effect and drug condition (none, MTP, PSIL) as a fixed effect. Left V1, right V1, and right A1 ROIs showed a significant effect of drug on activation. Post-hoc comparison (matlab: multcompare) for these 3 regions indicated that only left and right V1 showed differences in peak activation between non-drug and psilocybin conditions of p<0.05, uncorrected.
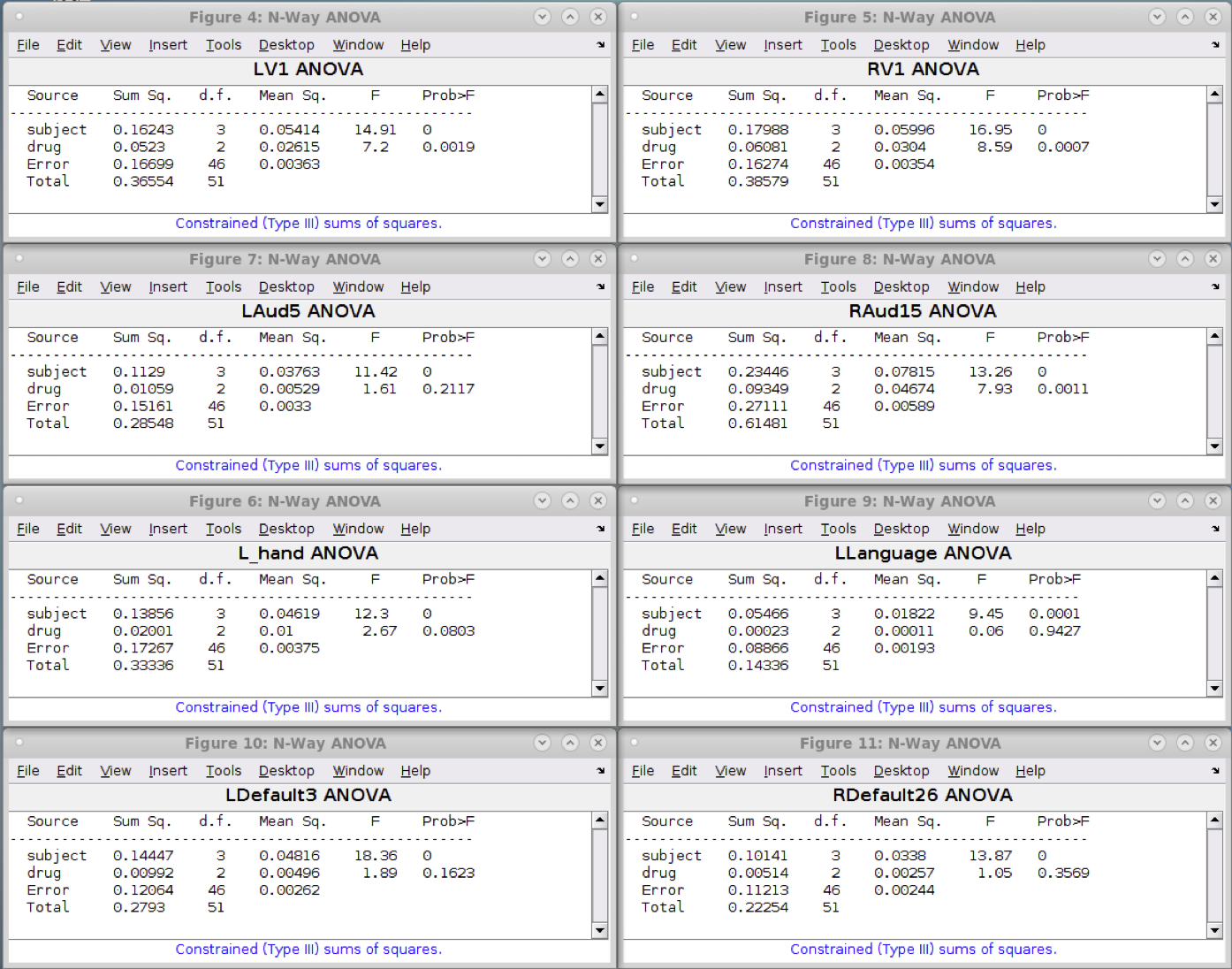
